# Mask-Based Breath Sampling for Detection of Pseudomonas aeruginosa in Adults with Cystic Fibrosis and Bronchiectasis

**DOI:** 10.64898/2026.06.14.26355606

**Authors:** Kasra Karimi, Harshitha Santosh Kumar, Sabine Wege, Katie Tiseo, Theresa Pfurtscheller, Elena Ivanova Reipold, Felix J Herth, Sabrina Klein, Ankur Gupta-Wright, Tobias Broger, Claudia M Denkinger

## Abstract

**Background:** Monitoring *Pseudomonas aeruginosa* (*P. aeruginosa*) infection in people with cystic fibrosis (pwCF) is essential for early detection, targeted treatment, and prevention of chronification. Sputum culture is current standard, yet many patients, particularly those receiving CFTR modulator therapy, struggle to expectorate sputum. Microbial aerosols from the respiratory tract offer a non-invasive alternative. This proof-of-principle study assessed the accuracy and feasibility of the AveloMask, a novel breath aerosol collection kit paired with qPCR detection.

**Methods:** Adult pwCF and bronchiectasis patients attending routine monitoring visits and healthy controls were enrolled in a cross-sectional study. Participants wore the mask for 30 minutes, followed by 20 instructed coughs. Mask filters were tested with a triplex qPCR assay targeting *P. aeruginosa* specific *ecfX* and *gyrB*, and human *RPP30* as an endogenous control. Accuracy was evaluated using a composite reference standard (sputum culture and PCR).

**Results:** Of 25 patients enrolled, 23 were included in the analyses. Sensitivity was 12/19 (63.2%) for breath qPCR versus 15/19 (78.9%) for sputum culture. Breath qPCR missed 5 cases detected by sputum culture but detected 2 sputum culture-negative/qPCR-positive cases. Specificity of breath qPCR was 100% in 4 patients and 15 healthy controls. *RPP30* was detected in all mask samples. AveloMask was perceived as easy to use, with many patients preferring it over sputum collection.

**Discussion:** Mask-based breath collection demonstrated promising diagnostic accuracy for detection of *P. aeruginosa*. Breath sampling may complement or partially substitute sputum-based diagnostics, especially in patients unable to expectorate. Further studies are needed to define its clinical role.

**HIGHLIGHTS:**

- Mask-based breath aerosol sampling with PCR detection demonstrated promising diagnostic performance for the detection of *P. aeruginosa* in adult people with Cystic Fibrosis (pwCF) and bronchiectasis patients.
- Breath-based PCR detected additional sputum culture-negative but sputum PCR-positive patients but also missed some sputum culture-positive cases.
- Breath sampling paired with molecular detection may complement sputum-based diagnostics, particularly for pwCF who cannot produce sputum, including those receiving CFTR modulator therapy.

## 1. Introduction

Cystic fibrosis (CF) typically causes chronic suppurative lung disease with recurrent infections and progressive respiratory failure [1]. Approximately 105,000 individuals are diagnosed with CF globally [2]. Chronic infection and colonization commonly occurs with *Pseudomonas aeruginosa* (*P. aeruginosa*) and is linked to a faster decline in lung function, more frequent exacerbations, and worse outcomes [3, 4]. Therefore, early detection and monitoring of respiratory tract bacteria are important parts of regular respiratory care [5].

Sputum culture is the standard method to identify lower respiratory tract pathogens in people living with CF (pwCF) [5]. However, some patients are unable to produce sputum, especially those on highly effective Cystic Fibrosis Transmembrane Conductance Regulator (CFTR) modulator therapy [6]. Sputum is commonly tested for pathogens using culture and [7] molecular detection, with comparable sensitivity, while some studies suggest that molecular detection might result in earlier disease detection [8]. Alternative methods, such as oropharyngeal swabs, can be used in patients unable to produce sputum, but show limited sensitivity in detecting lower airway pathogens [9]. Patients and clinicians see a strong need for non-invasive and easy to access sample types, with unmet need and minimal and optimal characteristics of tests having been described in a target product profile (TPP) [10]. In this context, breath sampling might offer an attractive alternative. Breath samples consist of small droplets or particles from the lower airways can contain microorganisms and microbial DNA [11]. Paired with PCR, this sampling technique has been evaluated in respiratory infections like tuberculosis and SARS-CoV-2, showing feasibility and high specificity in detecting pathogens [12, 13].

In this study, we investigated the use of a facemask breath sampling kit, AveloMask, for the detection of *P. aeruginosa* among pwCF and non-CF bronchiectasis. AveloMask has recently shown promising performance for PCR-based detection of *Mycobacterium tuberculosis,* and for detecting pathogens in patients with lower respiratory tract infections (LRTI) [14, 15]. When combined with quantitative PCR assays targeting *P. aeruginosa* –specific genes, this approach may offer a rapid and non-invasive method for pathogen detection in patients. The primary aim of this exploratory, proof-of-principle study was to evaluate the diagnostic accuracy of PCR performed on AveloMask-collected breath for detecting *P. aeruginosa* in adults with cystic fibrosis and non-cystic fibrosis bronchiectasis. Furthermore, usability and acceptability were assessed.

## 2. METHODS

### 2.1. Study Design

The study was conducted as a prospective, single-center, multi-gate cross-sectional diagnostic accuracy study at the Chest Hospital, Heidelberg University Hospital (Germany). The study was designed and reported in accordance with the Standards for Reporting Diagnostic Accuracy Studies (STARD) guidelines [16] (Table S1). Data collection methodology and analysis were planned before the commence of the study.

Adults (≥18 years) with a confirmed diagnosis of cystic fibrosis (CF) or non–cystic fibrosis bronchiectasis, were eligible, if they were scheduled to provide a sputum sample used for reference testing as part of routine clinical care. Exclusion criteria included:

- Initiation of anti-*Pseudomonas* antimicrobial therapy within 24 hours prior to enrollment.
- Use of inhaled antibiotics within 24 hours prior to the enrollment.
- Inability to provide informed consent.
- Not suitable to wear the AveloMask according to the study doctor due to underlying respiratory dysfunction.
- Failure to provide a sputum sample for routine microbiological testing.

Parallel to the main study, a group of healthy individuals were recruited to undergo the same sampling procedure to constitute a healthy control study population and evaluate specificity. All participants were asked to wear the AveloMask and resulting breath samples in buffer tubes and sputa were sent to Avelo for qPCR testing as described below.

### 2.2. Index Test

The index test aimed to detect *P. aeruginosa* DNA from breath samples collected using the AveloMask kit (Avelo AG, Switzerland). The kit includes a buffer tube (containing a nucleic acid-preserving solution) and a mask, similar to Filtering Face Piece 2 (FFP2) mask with an integrated filter inlay that can capture breath samples. The sample collection is done as follows (Figure 1 and Movie S1): Participants wore the mask for 30 minutes during normal tidal breathing and were instructed to perform 20 deep coughs at the end of the sampling period to increase aerosol yield. After wearing the mask (step 1), two stickers from each side of the mask, which protect the filter inlay during wear, are peeled off (step 2). The filter inlay was then pushed into the buffer tube using the stick attached to the tube cap (step 3). The buffer inactivates the sample and preserves nucleic acids for transport at ambient temperature until frozen for storage, extraction and molecular testing (step 4).

**Figure 1:**
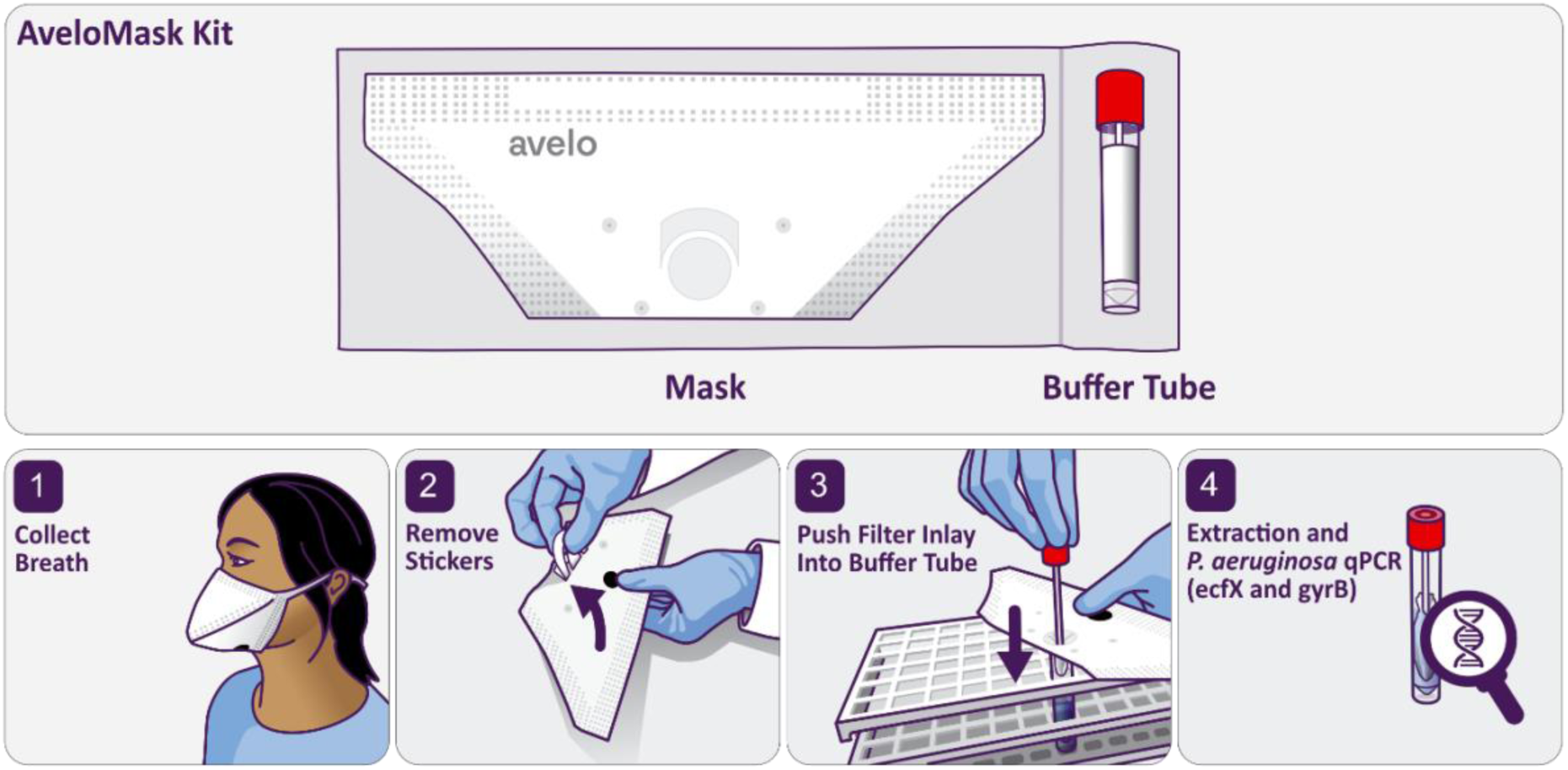
AveloMask Kit aerosol collection and testing procedure.

Breath samples were batch-tested using a triplex quantitative polymerase chain reaction (qPCR) assay, developed in-house, targeting *P. aeruginosa*–specific *ecfX* and *P. aeruginosa*–specific *gyrB* genes. Human RPP30 functioned as the endogenous control (Table S2). Both gene targets have previously been validated for specific detection of *P. aeruginosa* in respiratory specimens [17, 18]. A mask sample was considered positive if amplification of any bacterial gene target was detected within 40 cycles (Ct values). *P. aeruginosa* negative samples without amplification of the RPP30 endogenous control were considered invalid.

### 2.3. Reference tests

For patients in the chest hospital, a routine sputum sample was collected. Routine sputum culture for *P. aeruginosa* was performed in the diagnostic microbiology laboratory using standard methodology. Bacterial identification was performed by Matrix-Assisted Laser Desorption/Ionization Time-of-Flight (MALDI-TOF) mass spectrometry (see details in supplementary methods). All patients were asked to provide a second sputum sample and a subset of the patients was able to produce a second sputum. To minimize temporal bias, sputum collection and mask sampling were performed during the same outpatient visit. The additional sputum sample was evaluated with the same qPCR assay and same Ct cut-off for *P. aeruginosa* detection as the triplex qPCR used for breath sample testing [17, 18]. Additional clinical data such as other co-morbidities and information on respiratory performance metrics were also collected. Laboratory personnel performing breath qPCR analysis were blinded to sputum culture and sputum PCR results.

### 2.4. Acceptability and feasibility

Usability and acceptability were assessed using a brief post-sampling questionnaire. Patients rated the ease and comfort of AveloMask breath sampling on 5-point Likert scales, including comparison with sputum collection. The study physician documented procedural challenges, and open-ended questions collected qualitative feedback and suggestions for device improvement.

### 2.5. Statistical analysis

Sensitivity was assessed against a composite reference standard (CRS), in which the reference standard is defined as positive if either culture or qPCR tested positive from sputum, and negative if both were negative. Thus, discordant results (culture+/qPCR− or culture−/qPCR+) were classified as CRS-positive. Specificity was further assessed using AveloMask samples from healthy controls, assuming absence of infection in this group. For patients without sputum qPCR, CRS status was based on culture alone. Sample size was constrained by available patient volume at the study site and determined based on convenience. We targeted at least 15 *P. aeruginosa*-positive patients and an at least an equal number of reference-negative patients or healthy controls.

Diagnostic accuracy was evaluated comparing breath qPCR results with CRS of sputum culture and sputum qPCR. Two-sided 95% confidence intervals (CIs) were calculated using the Wilson score method [19]. Analyses were performed using R statistical software (version 4.4.1) [20]. A secondary objective was to assess the usability and acceptability of breath sampling using the AveloMask in interviews.

### 2.6. Ethical Approval

The study was conducted in accordance with the Declaration of Helsinki. Ethical approval was obtained from the Medical Faculty of Heidelberg Ethics Committee (approval number: S-778/2024). The study was registered on German clinical trials register [21], under the identification number “DRKS00036607” on 07.04.2025 prior to first enrollment. All participants provided written informed consent prior to enrollment.

## 3. RESULTS

### 3.1. Participant Flow and Study Population

A total of 25 patients and 15 healthy controls were enrolled between 07.04.2025 and 25.11.2025. Figure 2 summarizes the inclusion of the 25 patients enrolled during routine outpatient visits. Two patients were excluded post-enrollment: one due to recent inhaled antibiotic use (retrospectively found to violate inclusion criteria) and one due to AveloMask kit failure. Of the remaining 23 patients, all had index test and sputum culture results and 74% (17/23) had a second sputum sample for qPCR testing. All mask samples showed valid endogenous human RPP30 control amplification.

**Figure 2:**
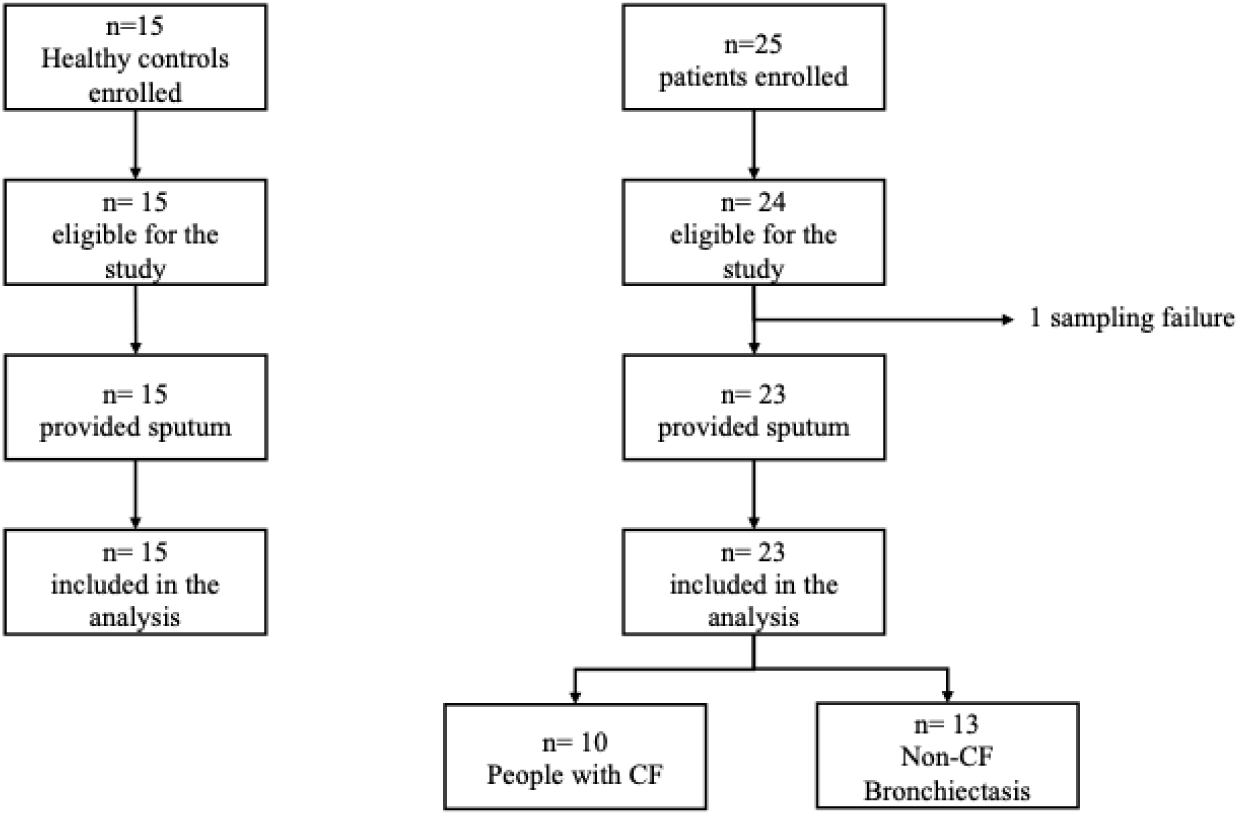
Flow Diagram. Note that due to different study procedure among enrolled patients and healthy controls, flow diagrams are demonstrated separately. Among the 25 patients enrolled, there was 1 enrollment failure due to post-enrolment disclosure of inhalative antibiotics usage within 24 hours prior to enrollment. CF: Cystic Fibrosis.

Baseline demographic and clinical characteristics are summarized in Table 1. Patients were adults with cystic fibrosis and non–cystic fibrosis bronchiectasis. The median age was 49, and 48% were female (11/23). Lung function, assessed by absolute forced expiratory volume in 1 s (FEV1), was 2.0 L at median. At time of enrollment, 13% were taking Azithromycin (3/23). The prevalence of *P. aeruginosa* positivity by sputum culture was 65% (15/23). Four patients were sputum qPCR positive. All of the enrolled pwCF were receiving CFTR modulator therapy (10/10).

**Table 1:** Baseline demographic and clinical characteristics of included patients for accuracy study.

| Characteristic | N=23 |
| --- | --- |
| Age in years | 49 (25 – 78) |
| Female sex at birth | 48% (11/23) |
| Body Mass Index | 24 (17 – 32) |
| **Primary respiratory diagnosis** |  |
| Cystic fibrosis | 43% (10) |
| Of those with Cystic fibrosis on current CFTR modulator use | 100% (10) |
| Non-CF bronchiectasis | 57% (13) |
| **Stable pulmonary health\*** | 70% (16) |
| FEV<sub>1</sub> | 2.0 L (0.78 L – 5.06 L) |
| **Treatment at enrollment** |  |
| Current oral Antibiotic use (azithromycin) | 13% (3) |
| **Sputum microbiology/culture at enrolment** |  |
| *P. aeruginosa* | 65% (15) |
| *Staphylococcus aureus* | 9% (2) |
| **Sputum *P. aeruginosa* culture density** |  |
| Low, 1+ | 57% (13) |
| Medium, 2+ | 4% (1) |
| High, 3+ | 4% (1) |
| Negative | 35% (8) |
| **Sputum *P. aeruginosa* qPCR** |  |
| Low, (Ct > 15) | 52% (12) |
| High (Ct ≤ 15) | 9% (2) |
| Negative | 13% (3) |
| Not done | 26% (6) |
| **Composite reference standard** |  |
| Positive culture or qPCR | 83% (19) |
| Negative culture and qPCR | 17% (4) |
| **Infection history** |  |
| History of *P. aeruginosa* | 100% (23) |

Data are presented as n% (n) or median (minimum-maximum).
FEV1: forced expiratory volume in 1 s; CFTR: cystic fibrosis transmembrane conductance regulator; History is defined as at least one positive culture of the pathogen within 12 months of screening;
\*Stable pulmonary health is based on judgement of the treating clinician.

### 3.2. Diagnostic Accuracy

Among the 19 CRS-positive patients, sensitivity of breath samples qPCR was 63.2% (12/19, 95% CI: 41.0%–80.9%) versus 78.9% (15/19, 95% CI: 56.7%–91.5%) for sputum culture (Table 2). Seven cases positive by the CRS were not detected by breath qPCR including 5 sputum culture+/qPCR+ cases and 2 culture-/qPCR+ cases. Conversely, breath qPCR detected 2 cases negative by sputum culture but positive by sputum qPCR.

**Table 2:**
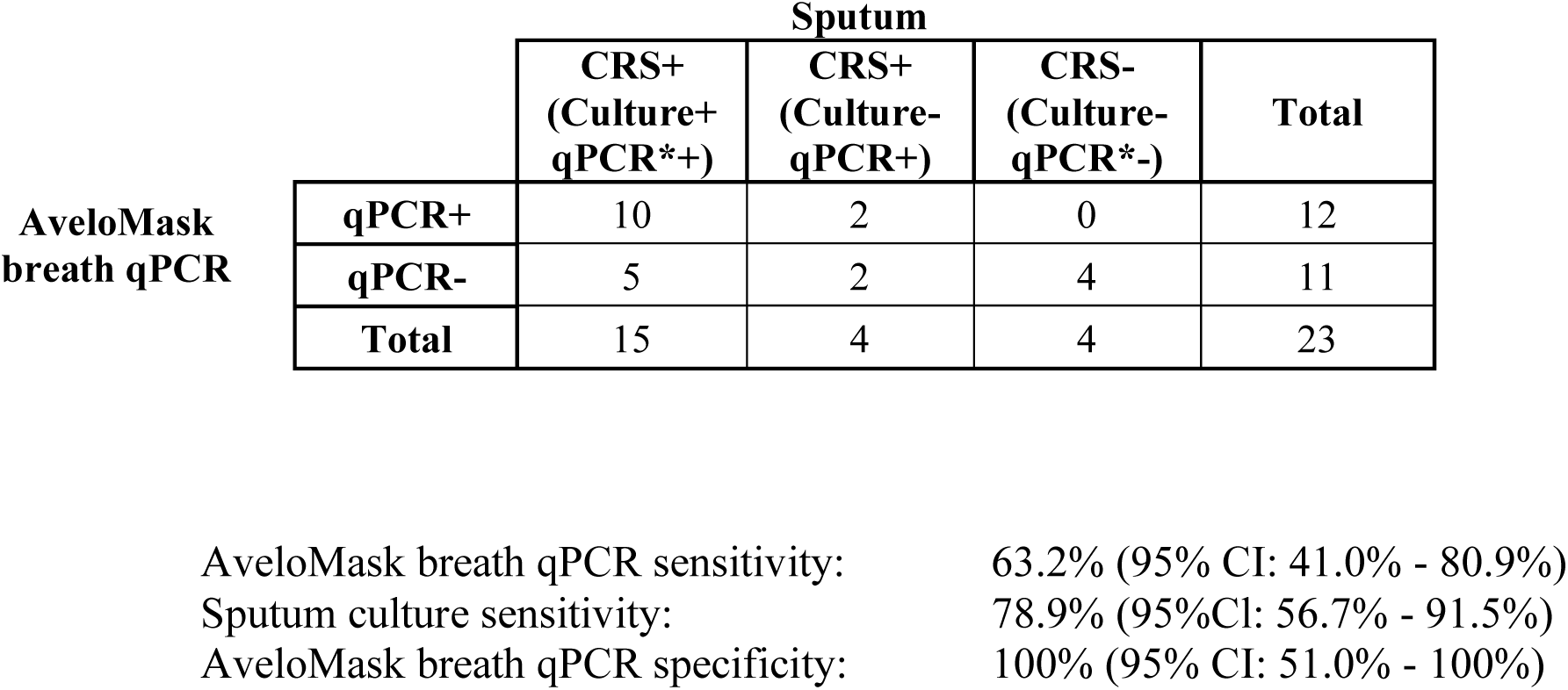
Contingency table comparing detection of *P. aeruginosa* using AveloMask breath qPCR with the composite reference standard (CRS) combining sputum culture and sputum qPCR. *Not all sputum samples have a qPCR result, in these cases, only the culture result is considered.

Among the 4 cases negative by the composite reference standard, breath qPCR correctly classified all 4 as negative, resulting in a specificity of 100% (95% CI: 51.0% - 100%). Consistently, in the healthy control group, all 15 samples were negative for the *P. aeruginosa*–specific targets, while the human RPP30 was detected in all samples (Table S3). Contingency tables using sputum culture (Table S4) and sputum qPCR (Table S5) as the reference standard show similar accuracies. Ct distribution of the *ecfX* and *gyrB* targets for breath and sputum are provided in the supplement (Figures S1 and S2).

### 3.3. Adverse Events

No adverse events related to mask sampling were reported. All participants (patients and healthy controls) were able to complete the study procedure.

### 3.4. Usability and Acceptability

For the assessment of usability and acceptability of the mask, the data from all 25 patients were included in the analysis (Figure 3A). According to the self-reported ease of providing the AveloMask sample, most patients found the procedure easy to perform. 48% (n=12) rated it as “Very easy,” and 32.0% (n=8) rated it as “Somewhat easy.” Only a small fraction, 8.0% (n=2), found the procedure “Somewhat difficult.” Overall, 80% (n=20) of the 25 respondents considered the process easy, indicating a high level of acceptability and ease of use among patients.

**Figure 3.**
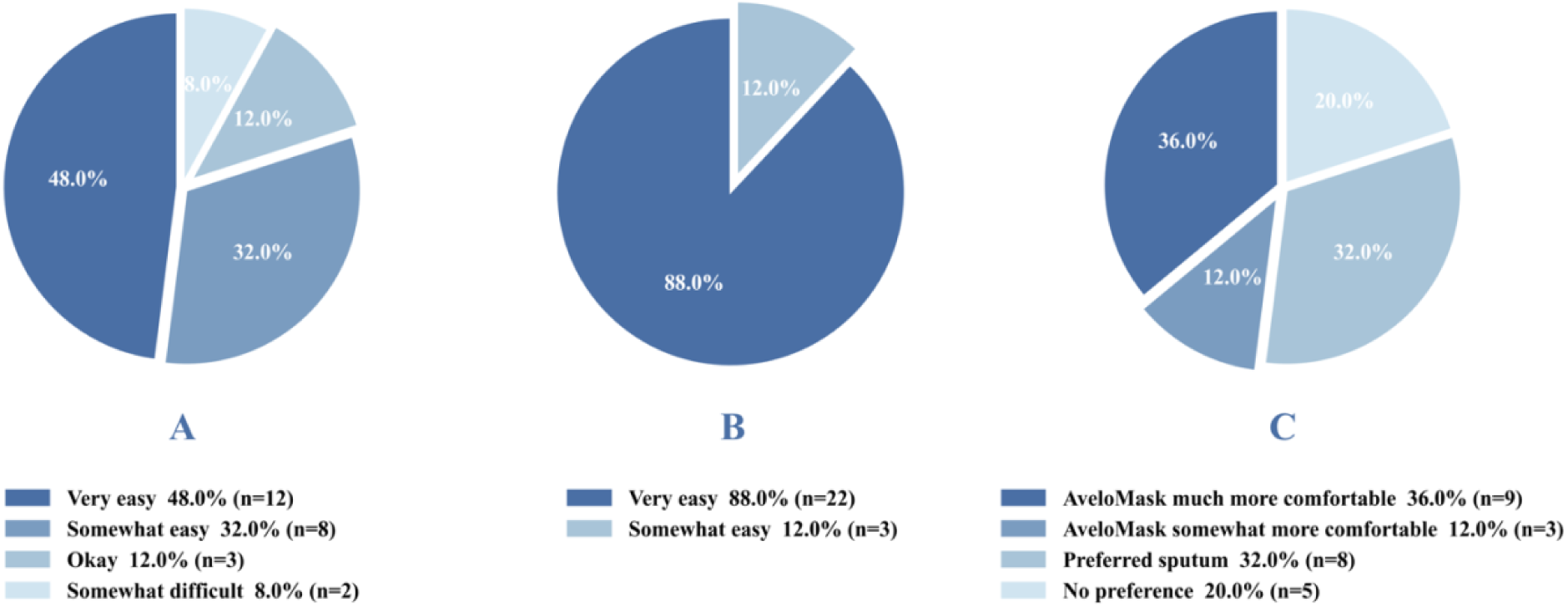
Usability and Acceptability Results. patients self-reported ease of use for providing the AveloMask breath sample (3A), study doctor ease of use assessment (3B), patient preference of AveloMask breath vs. sputum sampling (3C).

The assessing study doctor rated the AveloMask as easy to use for patients, with 88.0% (22/25) reporting it as “very easy” and 12.0% (3/25) as “somewhat easy” (Figure 3B). Any problems with the AveloMask sampling procedures were captured by the study doctor. The most frequent problem was difficulty transferring the filter inlay to the buffer tube (12.0%, 3/25). Other issues included the filter inlay detaching (4.0%, 1/25) and missing inlay attachment (4.0%, 1/25). The study doctor reported no issues with removing stickers. Patients provided various suggestions for improving the AveloMask (Supplemental Results).

The comfort rating of the AveloMask compared to sputum collection is detailed in Figure 3C. Among the 25 respondents, 36.0% (9/25) found the AveloMask breath collection to be much more comfortable than sputum collection. Additionally, 12.0% (3/25) felt that AveloMask breath collection was somewhat more comfortable than sputum collection. Meanwhile, 32.0% (8/25) preferred sputum collection, and 20.0% (5/25) had no preference, finding both methods equally comfortable.

## 4. DISCUSSION

Mask-based qPCR demonstrated moderate sensitivity and high specificity for detecting *P. aeruginosa* compared to sputum-based methods, accurately identifying many PCR-confirmed cases without false positives in PCR-negative patients and healthy individuals. Breath qPCR identified 12 CRS positive cases but missed 7 CRS positive cases (5 sputum culture+/qPCR+ and 2 culture-/qPCR+ cases), highlighting its potential as a non-invasive diagnostic approach while also indicating the need for further optimization to match the sensitivity of sputum-based testing.

The sensitivity of AveloMask in our study (63.2%) is lower than the 85% reported for dominant pathogen detection in pneumonia in Tiseo et al. [15]. This may be due to differences in aerosol generation between acute LRTI and chronic CF/non-CF bronchiectasis and likely reflects a combination of biological and methodological factors. Aerosol generation varies between individuals and depends on cough frequency, airway inflammation, and bacterial burden.

Overall, the sensitivity of mask-based sampling is lower than that of sputum-based sampling. While mask-based sampling enriches exhaled particles, the total biomass in aerosols is expected to be lower than in expectorated sputum. Optimization of DNA extraction protocols and assay sensitivity may further improve performance. Tiseo et al. [15] have further shown that the mask-based sampling preferably detects the dominant LRTI pathogen with reduced positivity for less dominant co-infections or colonisation. More studies are required to examine whether breath testing offers additional value in distinguishing clinically relevant infection from colonization, particularly if quantitative Ct values are correlated with clinical infection status and treatment decisions.

Sputum culture remains the clinical reference method for microbiological surveillance in CF, yet its sensitivity is limited and dependent on adequate sample expectoration [5]. Molecular techniques have consistently demonstrated increased pathogen detection compared with culture in respiratory samples [7, 18]. Our findings are consistent with this paradigm: discordant cases in which breath qPCR and sputum qPCR were positive despite negative culture likely reflect the higher analytical sensitivity of nucleic acid amplification testing. While we cannot exclude contamination through ‘dead’ *P. aeruginosa* DNA, all patients were known to have prior *P. aeruginosa* suggesting clinical relevance of the finding.

Sputum collection is increasingly difficult in patients on CFTR modulators, thus diagnostic yield may be an important complementary metric when evaluating breath-based sampling approaches. Diagnostic yield is defined as the proportion of people in whom the index test identifies *p. aeruginosa*, among *p. aeruginosa* positive patients for whom testing is undertaken. Although mask-based testing showed lower sensitivity than sputum-based molecular testing, the greater accessibility and availability of breath-based samples may partly compensate for this limitation by increasing opportunities for pathogen detection and longitudinal microbiological surveillance in patients unable to reliably expectorate sputum. The concept of diagnostic yield has been well described for tuberculosis [22].

Consistent with the Tiseo et al. study, patients in our cohort found the mask easy to use and tolerated it well, suggesting its feasibility for in- and outpatient settings across different patient populations, including people with suspected pneumonia and cystic fibrosis.

Strengths of the study include the use of a quality assured AveloMask sampling kit (produced under a quality management system) and a well-described triplex qPCR test targeting two widely used *P. aeruginosa* genes, the use of human endogenous controls, enrollment of pwCF and non-CF bronchiectasis patients in a real-world setting and a CRS combining sputum culture and qPCR. Several limitations must be acknowledged: (1) Due to the small sample size, confidence intervals for specificity are wide; (2) healthy controls were included because only few CRS-negative patients were available; and (3) some patients did not have sputum-PCR results. (4) The CRS might lead to incorporation bias but analyses with alternative reference standards were presented. (5) Furthermore, the ability to produce sputum was designated as an inclusion criterion. While this design choice was necessary to enable comparison with the reference standard, it restricts generalizability to the population most likely to benefit from mask-based sampling, namely individuals unable to expectorate sputum. (6) The study was conducted at a single tertiary center, and performance may differ in other clinical settings or patient populations. Finally, (7) antimicrobial resistance profiles were not assessed, limiting insights into the clinical relevance of detected strains.

Future research should include patients unable to produce sputum to evaluate the diagnostic yield and clinical utility of mask-based testing, the patient group who might benefit the most from an alternative sampling procedure. Furthermore, direct comparison of mask sampling with oropharyngeal swabs and induced sputum and testing of additional pathogens will be necessary. Also, the population in our study includes adults with established chronic airway disease under routine clinical monitoring at a specialized center, thus results may not directly generalize to other populations, e.g. children, early-stage disease, or acute exacerbations, which should be included in future studies. Additionally, we did not assess inter-operator variability or repeatability of mask sampling, which are important considerations for broader implementation. Further multicenter studies with larger and more diverse cohorts are required. On the analytical side, future research should focus on improving DNA extraction methods and integrating them with commercial molecular multiplex testing platforms to enhance sensitivity, cover clinically relevant pathogens and broaden clinical utility.

Mask-based breath aerosol PCR demonstrated moderate sensitivity and high specificity for detection of *P. aeruginosa* in adults with cystic fibrosis and bronchiectasis. The method was feasible, well tolerated, and capable of identifying positive cases not detected by culture. While current performance does not support replacement of sputum diagnostics, aerosol-based molecular testing represents a promising non-invasive addition for respiratory pathogen surveillance, especially in the patient group that cannot produce sputum. Larger, adequately powered studies are required to refine performance estimates and further define the role of breath qPCR in clinical practice.

## DECLARATIONS

### Presentation

The project was presented as a poster at DZIF/PEG joint meeting in February 2026[23], and at ESCMID Global[24] in April 2026.

### Availability of data and materials

All data generated or analysed during this study are included in this published article, in the supplemental material, and supplemental data spreadsheet.

### Credit authorship contribution statement

Authorship order follows contribution and ICMJE criteria.

## Acknowledgements

The authors thank the study participants and clinical staff, especially the Clinical team at Thorax clinic for their contributions. This research was supported Innosuisse (award: 107.660 SIP-LS) and by Avelo Inc. (Switzerland). Avelo staff was involved in the design, data collection, analysis and decision to publish.

## Declaration of generative AI in scientific writing

During the preparation of this work the authors used Mistral.AI to draft and proof-read the text. After using this tool/service, the authors reviewed and edited the content as needed and take full responsibility for the content of the published article.

## ANNEX

### Supplementary materials

**Table S1:** STARD Checklist.

| Section & Topic | No | Item | Done? |
| --- | --- | --- | --- |
| **TITLE OR ABSTRACT** |  |  |  |
|  | **1** | Identification as a study of diagnostic accuracy using at least one measure of accuracy (such as sensitivity, specificity, predictive values, or AUC) | yes |
| **ABSTRACT** |  |  |  |
|  | **2** | Structured summary of study design, methods, results, and conclusions (for specific guidance, see STARD for Abstracts) | yes |
| **INTRODUCTION** |  |  |  |
|  | **3** | Scientific and clinical background, including the intended use and clinical role of the index test | yes |
|  | **4** | Study objectives and hypotheses | yes |
| **METHODS** |  |  |  |
| *Study design* | **5** | Whether data collection was planned before the index test and reference standard were performed (prospective study) or after (retrospective study) | yes |
| *Participants* | **6** | Eligibility criteria | yes |
|  | **7** | On what basis potentially eligible participants were identified (such as symptoms, results from previous tests, inclusion in registry) | yes |
|  | **8** | Where and when potentially eligible participants were identified (setting, location and dates) | yes |
|  | **9** | Whether participants formed a consecutive, random or convenience series | yes |
| *Test methods* | **10a** | Index test, in sufficient detail to allow replication | yes |
|  | **10b** | Reference standard, in sufficient detail to allow replication | yes |
|  | **11** | Rationale for choosing the reference standard (if alternatives exist) | yes |
|  | **12a** | Definition of and rationale for test positivity cut-offs or result categories of the index test, distinguishing pre-specified from exploratory | yes |
|  | **12b** | Definition of and rationale for test positivity cut-offs or result categories of the reference standard, distinguishing pre-specified from exploratory | yes |
|  | **13a** | Whether clinical information and reference standard results were available to the performers/readers of the index test | yes |
|  | **13b** | Whether clinical information and index test results were available to the assessors of the reference standard | yes |
| *Analysis* | **14** | Methods for estimating or comparing measures of diagnostic accuracy | yes |
|  | **15** | How indeterminate index test or reference standard results were handled | yes |
|  | **16** | How missing data on the index test and reference standard were handled | yes |
|  | **17** | Any analyses of variability in diagnostic accuracy, distinguishing pre-specified from exploratory | no |
|  | **18** | Intended sample size and how it was determined | no |
| **RESULTS** |  |  |  |
| *Participants* | **19** | Flow of participants, using a diagram | yes |
|  | **20** | Baseline demographic and clinical characteristics of participants | yes |
|  | **21a** | Distribution of severity of disease in those with the target condition | yes |
|  | **21b** | Distribution of alternative diagnoses in those without the target condition | no |
|  | **22** | Time interval and any clinical interventions between index test and reference standard | yes |
| *Test results* | **23** | Cross tabulation of the index test results (or their distribution) by the results of the reference standard | yes |
|  | **24** | Estimates of diagnostic accuracy and their precision (such as 95% confidence intervals) | yes |
|  | **25** | Any adverse events from performing the index test or the reference standard | yes |
| **DISCUSSION** |  |  |  |
|  | **26** | Study limitations, including sources of potential bias, statistical uncertainty, and generalisability | yes |
|  | **27** | Implications for practice, including the intended use and clinical role of the index test | yes |
| **OTHER INFORMATION** |  |  |  |
|  | **28** | Registration number and name of registry | yes |
|  | **29** | Where the full study protocol can be accessed | no |
|  | **30** | Sources of funding and other support; role of funders | yes |

**Table S2:**
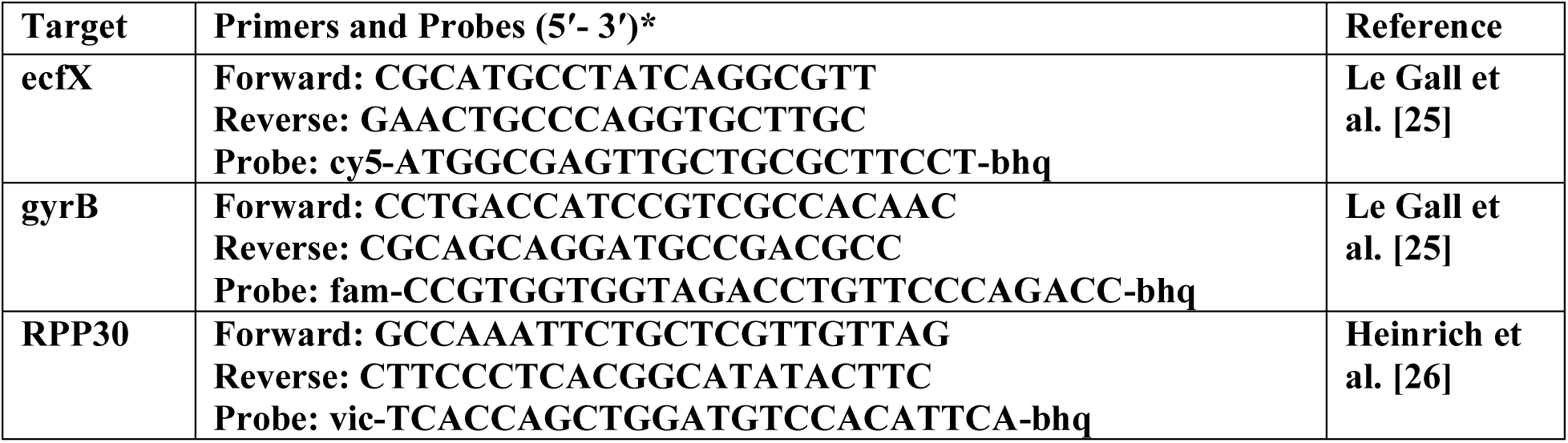
PCR Primers and Probes. *.

**Table S3:** Contingency table for comparing qPCR results of different target sequences of the healthy negative control group.

|  | AveloMask<br>breath<br>qPCR |  |  |
| --- | --- | --- | --- |
|  | + | - | Total |
| PCR<br>Target<br>Gene |  |  |  |
| ecfX | 0 | 15 | 15 |
| gyrB | 0 | 15 | 15 |
| RPP30 | 15 | 0 | 15 |

**Table S4:** Contingency table comparing detection of *P. aeruginosa* using AveloMask breath qPCR with sputum culture as reference.

|  | Sputum Culture |  |  |
| --- | --- | --- | --- |
|  | + | - | Total |
| AveloMask<br>breath qPCR |  |  |  |
| + | 10 | 2 | 12 |
| - | 5 | 6 | 11 |
| Total | 15 | 8 | 23 |

**Sensitivity:** 66.7% (95% CI: 41.7% - 84.8%)
**Specificity:** 75.0% (95% CI: 40.9% - 92.9%)

**Table S5:** Contingency table comparing detection of *P. aeruginosa* using AveloMask breath qPCR with sputum qPCR as reference.

|  | Sputum PCR |  |  |
| --- | --- | --- | --- |
|  | + | - | Total |
| AveloMask<br>breath qPCR |  |  |  |
| + | 9 | 0 | 9 |
| - | 5 | 3 | 8 |
| Total | 14 | 3 | 17 |

**Sensitivity:** 64.3% (95% CI: 38.8% - 83.7%)
**Specificity:** 100% (95% CI: 43.9% - 100%)

### Supplementary Methodic - bacteriological reference standard

Viscous sputum was treated with liquilizer (Meta Systems) according to the manufacturer’s instructions before inoculation of the media. Standard cultivation was done with Columbia blood sheep agar (BD Diagnostics), chocolate agar, Macconkey Agar, (BioMérieux) Sabouraud-Glucose-Agar, Burkholderia cepacian agar (BCSA) and CandidaChrom Agar (BD) for at least 48h at 36+/-1°C and 5% CO2, with cultivation of BCSA at 30+/-1°C and incubation of Sabouraud agar and BCSA for at least 7 days. For identification, MALDI-TOF (Bruker Corporation, Billerica, MA) was used.

### Supplementary Results - Usability – Improvement ideas

The most common suggestion was to reduce the time needed (40.0%, 10/25), followed by changing the design (24.0%, 6/25). Other suggestions included reducing the number of coughs required (16.0%, 4/25), making the mask more breathable (12.0%, 3/25), and removing the cough requirement entirely (4.0%, 1/25). Notably, 28.0% (7/25) of patients felt that no improvements were needed.

**Figure S1.**
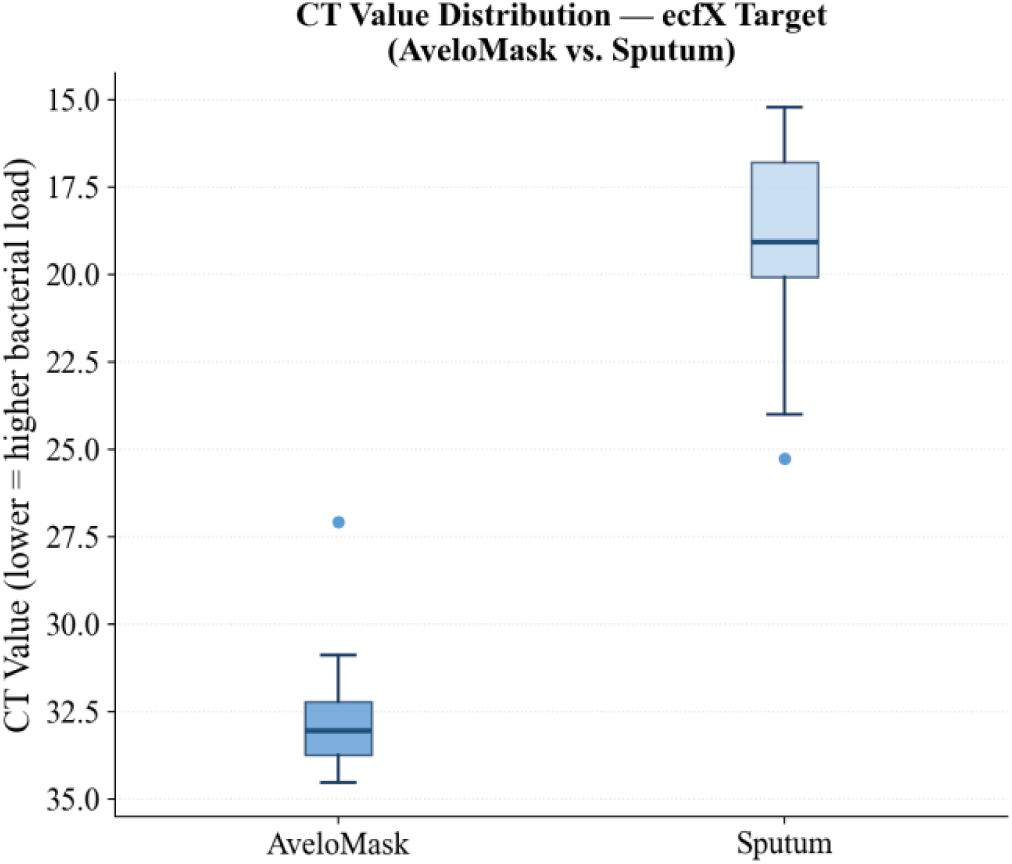
ecfX CT distributions — AveloMask vs. Sputum

**Figure S2.**
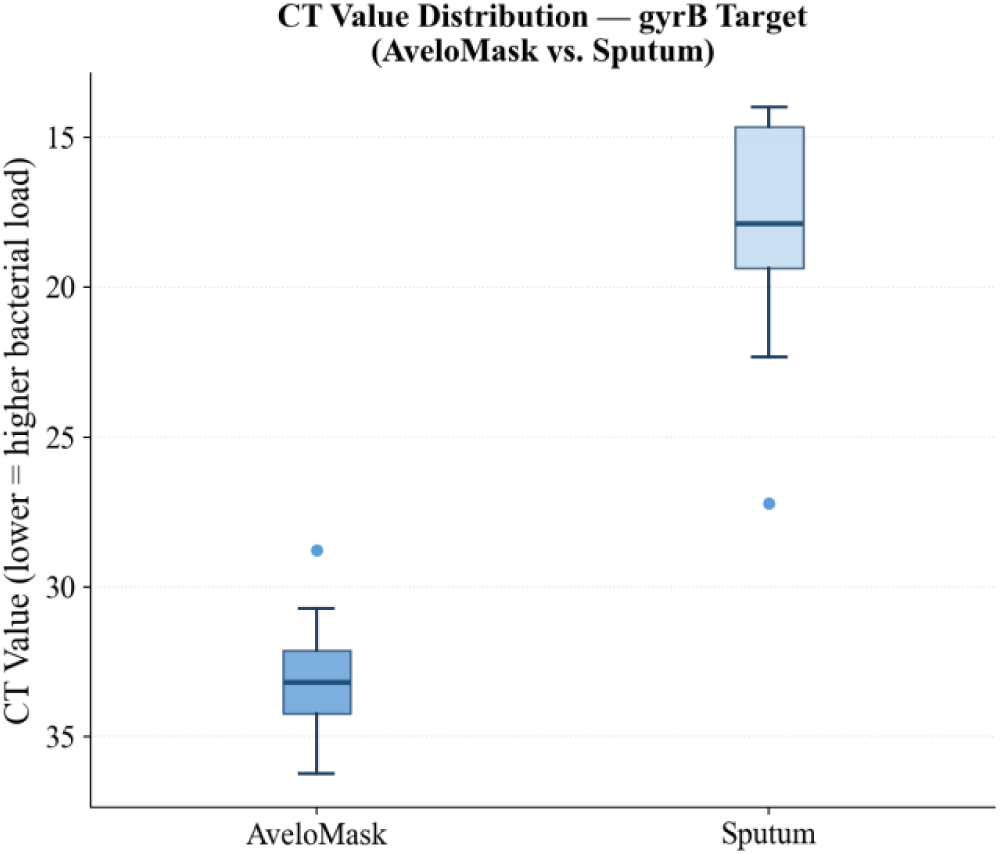
gyrB CT distributions — AveloMask vs. Sputum.

